# Bayesian networks to estimate prognosis in vascular cognitive impairment and small vessel disease: integrated analyses of interdependent contributors to multiple outcomes

**DOI:** 10.64898/2026.06.03.26354793

**Authors:** L. Malin Overmars, Cor Allaart, Esther E. Bron, Hans-Peter Brunner La Rocca, Jeroen de Bresser, Majon Muller, Matthias J.P. van Osch, Charlotte Teunissen, Betty M Tijms, Frank J. Wolters, Geert Jan Biessels, the Heart-Brain Connection Consortium

## Abstract

**Background:** Vascular cognitive impairment (VCI) and small vessel disease (SVD) involve many interconnected factors influencing multiple outcomes, also beyond cognitive decline. Bayesian networks (BNs) can help unravel these complex interrelations, which we demonstrate in this proof-of-concept study in the Heart-Brain Connection cohort, including memory-clinic patients with SVD, patients with heart failure, carotid occlusive disease, and reference participants.

**Methods:** We trained BNs and jointly modelled cognitive decline (Clinical Dementia Rating (CDR) increase) and major adverse cardiovascular events (MACE) over five years as outcomes in relation to multiple demographic and disease factors and emerging imaging and plasma biomarkers, also considering possible non-random dropout.

**Results:** Of 566 individuals (median age 68, 64% men), 134 had MACE and 112 experienced CDR increase. Diagnostic group and baseline cognition were key determinants of both outcomes. The BN identified baseline clinical severity as a non-random dropout source. Plasma biomarkers formed an interconnected subnetwork, linked to demographic and vascular factors, but without direct dependencies with outcomes. The trained BN also provides individualized inference under partial evidence, informing on outcome probabilities.

**Conclusion:** This proof-of-concept study demonstrates how BNs quantify and visualize the dependency structure underlying prognostic heterogeneity in VCI and SVD, including non-random dropout and positioning of emerging biomarkers.

## Introduction

Vascular cognitive impairment (VCI) and small vessel disease (SVD) are heterogeneous constructs with inherent variability in vascular risk profiles and etiologies, clinical symptoms, and long-term outcomes (1). The latter include cognitive decline, but also adverse cardiovascular events. Estimating which individuals will experience these outcomes is challenging, because the many contributing risk and disease factors are interconnected in complex, often non-linear ‘networks’ and may influence adverse outcomes directly and indirectly. An additional challenge is that substantial numbers of patients may drop out from research cohorts, due to the conditions themselves, but also due to advanced age and comorbidities, leading to attrition bias (2–4). Finally, with an increasing number of specialized diagnostic and prognostic plasma biomarkers becoming available, it remains unclear which markers have additional value over clinical predictors for improving prognostic models.

A network-based approach that jointly models and weighs interrelated factors across multiple outcomes, while keeping the relationships transparent, can help address several recurring challenges in prognostic studies in VCI and SVD (e.g., correlated predictors, heterogeneous patient profiles, and incomplete follow-up) (5–7). Here, Bayesian networks (BNs) are of particular interest. Although the method has existed for decades, its use in healthcare has accelerated in recent years, driven by increasing data availability and computational power (8). BNs represent factors considered as a directed graph of conditional dependencies, revealing which factors are (in)directly linked, and how combinations of factors jointly shape outcome probabilities (9). Because BNs represent the conditional dependencies among all variables, they can use observed patient information to infer the probable states of unobserved variables through the network, enabling risk estimation from incomplete data. Also, BNs can support individualized and explainable risk estimation and “what-if” scenarios (e.g., how predicted risks shift under different constellations of vascular risk, cognitive performance, and existing brain injury), as shown in prior dementia and post-stroke prediction applications (5–8,10–12).

Here, we show proof-of-concept of the potential of BNs in unraveling, visualizing, and quantifying complex interrelations of disease factors and outcomes in VCI and SVD, using the Heart-Brain Connection (HBC) cohort (13). HBC was designed to assess the role of hemodynamic factors in VCI, in patient populations with different cardiovascular conditions. These include heart failure (HF), carotid occlusive disease (COD), and memory clinic patients with vascular brain lesions, predominantly SVD, conditions with shared and distinct factors affecting the brain. A reference group was also included. In HBC, standardized data on demographics, vascular risk factors, disease characteristics, brain MRI, functional status, and 5-year clinical outcomes are collected. Cognitive impairment, at baseline and during follow-up, was monitored in all three groups and a reference group (14). This design makes HBC particularly well-suited for our BN concept study, because of the standardized, detailed work-up and outcome assessment, combined with the heterogeneity and complexity of underlying diseases in the different patient groups. We present three showcases. First, we demonstrate how a BN can jointly model interdependencies in factors related to two key clinical outcomes, cognitive decline and cardiovascular events, as well as study dropout, and dropout reasons. Second, we examine how emerging imaging and plasma biomarkers can be included in the network and how they connect to established risk factors, imaging and outcomes, and, lastly, we show how probabilistic inference can be translated to the individual patient level by calculating posterior probabilities based on new evidence.

## Material and Methods

### 2.1 Study population

We used data from the HBC study, a multicenter, observational study (13). In-person and telephone follow-up was done after two and five years. Inclusion criteria for all groups were age ≥ 50 years, ability to undergo cognitive testing, and independence in daily life. Exclusion criteria were contraindications for MRI or inability to complete the MRI protocol, life-threatening disease with life expectancy < 3 years (other than VCI, COD, or HF), neurodegenerative disease other than VCI or Alzheimer’s disease, other neurological or psychiatric disorders that could affect cognition (e.g. severe traumatic brain injury, substance abuse), participation in therapeutic intervention trials, planned relocation outside the region within 3 years, and clinically relevant arrhythmia at inclusion (atrial fibrillation or frequent premature ventricular contractions).

Patients with possible VCI additionally required cognitive complaints, a Clinical Dementia Rating (CDR) ≤ 1 and Mini-Mental State Examination (MMSE) ≥ 20, and evidence of vascular brain injury: either moderate–severe white matter hyperintensities (Fazekas > 1), lacunes and/or intracerebral (micro)hemorrhages, or mild white matter lesions (Fazekas = 1) in combination with at least two vascular risk factors (hypertension, hypercholesterolemia, diabetes mellitus, obesity, smoking, or clinically manifest vascular disease). COD patients were required to have > 80% stenosis or occlusion of the internal carotid artery on MR angiography and not be scheduled for carotid surgery. HF patients fulfilled European Society of Cardiology criteria for HF, with typical symptoms and signs, objective cardiac dysfunction on echocardiography, and a clinically stable condition for ≥ 6 months. Reference participants were recruited through advertising leaflets and among spouses of patients.

### 2.2 Outcome definitions

The first outcome considered, cognitive decline, was defined as either institutionalization in a nursing or care home at year 2 or year 5, or an increase of at least 0.5 points in CDR global score over time (15). These outcomes were recorded during follow-up through in-person assessment or telephone interview at year 2, and through (informant) telephone interview at year 5 using standardized interviews. The second outcome, major adverse cardiovascular events (MACE), was defined as cardiac death, cerebrovascular accident (including transient ischemic attack, intracerebral hemorrhage, and stroke), myocardial infarction, angioplasty/stent placement, coronary artery bypass surgery, surgery or stent placement for carotid stenosis, and hospital admission for HF, as recorded at follow-up visits or by phone at year 2 and by phone in year 5 through standardized interviews (16). When no information on MACE was available at one or both time points, we defined the outcome as unobserved. For deceased participants, the primary cause of death was obtained from Central Agency for Statistics Netherlands (CBS) and general practitioners, if known.

### 2.3 Variable selection, definitions, and processing

The variable selection for network creation was guided by clinical domain knowledge to balance clinical relevance with the need for stable probability estimation. In a BN, each variable and its categories increase the number of conditional probability tables (CPT) that must be estimated, and a moderate sample size limits the number of variables that can be reliably modeled. We therefore selected a focused set of clinically relevant variables covering demographics (age, sex), cardiovascular risk (SCORE2/SCORE2-OP) (17,18), imaging markers of cerebrovascular damage (SVD score, total brain volume, hippocampal volume), cardiovascular disease history (atherosclerotic cardiovascular disease, cerebrovascular accident history, patient group), baseline cognitive and functional status (CDR, MMSE, Starkstein depression score).

For the second showcase, the network was extended with cerebral blood flow (CBF), emerging plasma biomarkers of neurodegenerative disease (Amyloid beta 42 (Aβ42), Amyloid beta 40 (Aβ40), glial fibrillary acidic protein (GFAP), neurofilament light chain (NfL), phosphorylated tau 181 (pTau181)) and two previously published plasma biomarker compound scores (BCS) reflecting inflammation and coagulation (19–21).

We derived all BN variables from the harmonized baseline and follow-up HBC dataset and performed several preprocessing steps (public code: https://github.com/umcu/VCI-Bayes). Age and sex were taken from baseline records. Vascular risk was summarized using the SCORE2 and SCORE2-OP algorithms from the European Society of Cardiology, which estimates 10-year risk of atherosclerotic cardiovascular disease based on age, sex, smoking status, systolic blood pressure, diabetes, total cholesterol and high-density lipoprotein cholesterol (17,18,22). History of atherosclerotic cardiovascular disease was defined as any prior ischemic stroke or transient ischemic attack (TIA), myocardial infarction, percutaneous coronary intervention or coronary bypass surgery, or peripheral arterial disease at baseline. Brain volume measures were expressed as a percentage of intracranial volume. Baseline cognition and mood were captured by the MMSE total score, the global CDR, and the Starkstein depression score. SVD-burden was summarized using a neuroradiological SVD score, based on scores/presence for white matter hyperintensities (Fazekas), lacunar infarcts, perivascular spaces, and microbleeds (23). More details regarding MRI processing and plasma biomarker measurements can be found in the Supplementary Methods.

### 2.4 BN training

For the following analyses, we used pyAgrum (version 2.2.0), a Python wrapper and library built upon the C++ aGrUM framework (24,25). The pyAgrum library is dedicated to BNs and other probabilistic graphical models.

#### 2.4.1 Variable discretization and algorithm

All continuous variables were discretized into 4 quartile-based bins using PyAgrum’s DiscreteTypeProcessor (24,25). To learn the network structure, we employed the K2 algorithm, a score-based search method implemented in PyAgrum (24,25). We selected K2 specifically for its ability to integrate data-driven learning with domain knowledge through predefined topological ordering, as shown in the next paragraph. This allowed us to strictly enforce the clinical hierarchy described below, preventing implausible causal directions (e.g., outcomes influencing baseline characteristics). Mechanistically, the algorithm iterates through the ordered variables and, for each node, greedily selects the set of parent variables from preceding layers that maximizes the network’s Bayesian score. The scoring function approximates the marginal likelihood of the data given the structure, utilizing a Dirichlet prior with uninformative hyperparameters to penalize overly complex graphs. This approach balances model fit with parsimony, ensuring that the resulting network captures robust conditional dependencies while minimizing the risk of overfitting inherent in moderate sample sizes.

#### 2.4.2 Clinically informed network structure

The variable order within the network was imposed by means of a clinically informed structure to reflect a potential causal order. The layered hierarchy constrained the search space by allowing directed edges only from higher to lower layers or within the same layer, thereby preventing implausible directions (e.g., occurrence of stroke pointing towards age). The functions addForbiddenArc() and addMandatoryArc() were used to implement these constraints. The hierarchy was defined by MO and GJB based on prior knowledge and clinical reasoning, as follows:

#### At inclusion in the study

**Layer 0** – Unmodifiable demographics (age, sex)

**Layer 1** – Cardiovascular risk factors (vascular risk score - SCORE2 and SCORE2-OP)

**Layer 2** – Emerging imaging and plasma biomarkers (CBF, Aβ42, Aβ40, GFAP, NfL, pTau181, and two previously published Olink plasma BCS reflecting inflammation and coagulation (19,20)) – added in second analysis, see below

**Layer 3** – Imaging markers of neurovascular damage (SVD score, total brain volume/intracranial volume, and hippocampal volume/intracranial volume), under the assumption that these may precede the clinical diagnoses in the next layers

**Layer 4** – Current and previous cardiovascular diagnoses (atherosclerotic cardiovascular disease history, cerebrovascular accident history, patient group)

**Layer 5** – Cognitive status (baseline CDR, MMSE), under the assumption that all the preceding layers may affect cognition, rather than vice versa

#### During follow-up

**Layer 6** – Outcomes (CDR increase, MACE, Unobserved)

**Layer 7** – Dropout reason

For comparison, an unconstrained network was learned, in which no hierarchical restrictions were applied. This illustrates the influence of the layer constraints on the learned structure and demonstrates the extent to which unconstrained learning may produce non-plausible relationships.

To evaluate the robustness of the learned network structure, a nonparametric bootstrap analysis was performed. The original dataset was resampled 200 times with replacement, maintaining the same sample size in each iteration. For each bootstrap replicate, a new BN was learned. Across replicates, the presence or absence of each directed edge was recorded, and the relative frequency of each arc was computed.

### 2.5 Analyses

Using the learned network structure, we performed three analyses.

#### 2.5.1 Jointly modelling cognitive decline, cardiovascular events, and dropout

To inspect how the jointly modelled interdependencies among factors, outcomes, and dropout relate to the outcome nodes, we examined the structure and conditional probability tables (CPT) of the two outcome nodes in the learned BN. In a BN, each node stores a CPT that specifies the probability distribution of that variable conditional on all possible configurations of its parent variables. Inspecting these tables therefore shows how the model assigns outcome probabilities under different combinations of associated factors.

#### 2.5.2 Positioning of emerging imaging and plasma biomarkers within the network

To examine how emerging imaging and plasma biomarkers are positioned within the network, we added an additional layer to the network: L2 – Emerging imaging and plasma biomarkers of neurodegenerative disease, i.e. CBF, Aβ42, Aβ40, GFAP, NfL, pTau181, and two previously published Olink plasma BCS reflecting inflammation and coagulation (19,20). We placed this layer between cardiovascular risk factors and imaging markers of neurovascular damage and again learned 200 bootstrapped BNs, after which the presence or absence of each directed edge was recorded, and the relative frequency of each arc was computed. Based on the location of the biomarkers in the network and the incoming and outgoing arcs, we determined their dependency relationship with respect to the outcomes.

To provide a known reference point, we also applied standard logistic regression models. These models estimate the association between each biomarker and both outcomes (CDR increase and MACE), both unadjusted and adjusted for age and sex. The logistic regression evaluates the biomarkers independently of the vascular, imaging, and functional factors that are jointly modeled in the network. Including these analyses clarifies whether associations that emerge in logistic regression persist or disappear when the broader dependency structure is considered.

#### 2.5.3 Probabilistic inference at the individual patient level

We illustrated probabilistic inference at the individual level in two complementary ways. First, we defined a single prototypical profile representing a cognitively healthy individual with low vascular risk at baseline: a 58-year-old female from the reference group, with an SVD score of 0, MMSE score of 29, and baseline CDR of 0. All other variables were left unobserved. The model generated individualized posterior probabilities over cognitive decline (yes, no, unobserved) and MACE (yes, no, unobserved), conditional on this limited evidence. To assess the stability of these estimates, we repeated the inference procedure across 200 bootstrap networks. For each bootstrap sample, a new Bayesian network was learned, posterior probabilities for this profile were recalculated, and the resulting distributions were summarized. From these 200 draws, we derived the mean, standard deviation, and 95% confidence intervals for each outcome category. Second, to illustrate how one or more factors in the network can be ‘tweaked’ and how the posterior probabilities update accordingly, we set the evidence to fixed values (age 72, female), and visualized how posterior probabilities of both outcomes change as the SVD score increases.

## Results

### Baseline characteristics

A total of 566 participants were included (Table 1). The median age was 68 years (IQR 62–74), and 364 (64%) were men. The cohort comprised 162 (29%) patients with HF, 166 (29%) with VCI from memory clinics, 109 (19%) with COD, and 129 (23%) reference participants. Of all 566 individuals, 134 had MACE during follow-up and 112 had an increase in CDR, 39 individuals experienced both. Of 213 individuals, no complete 5-year outcomes were observed. Reasons for study discontinuation per patient group are shown in Figure 1.

**Figure 1.**
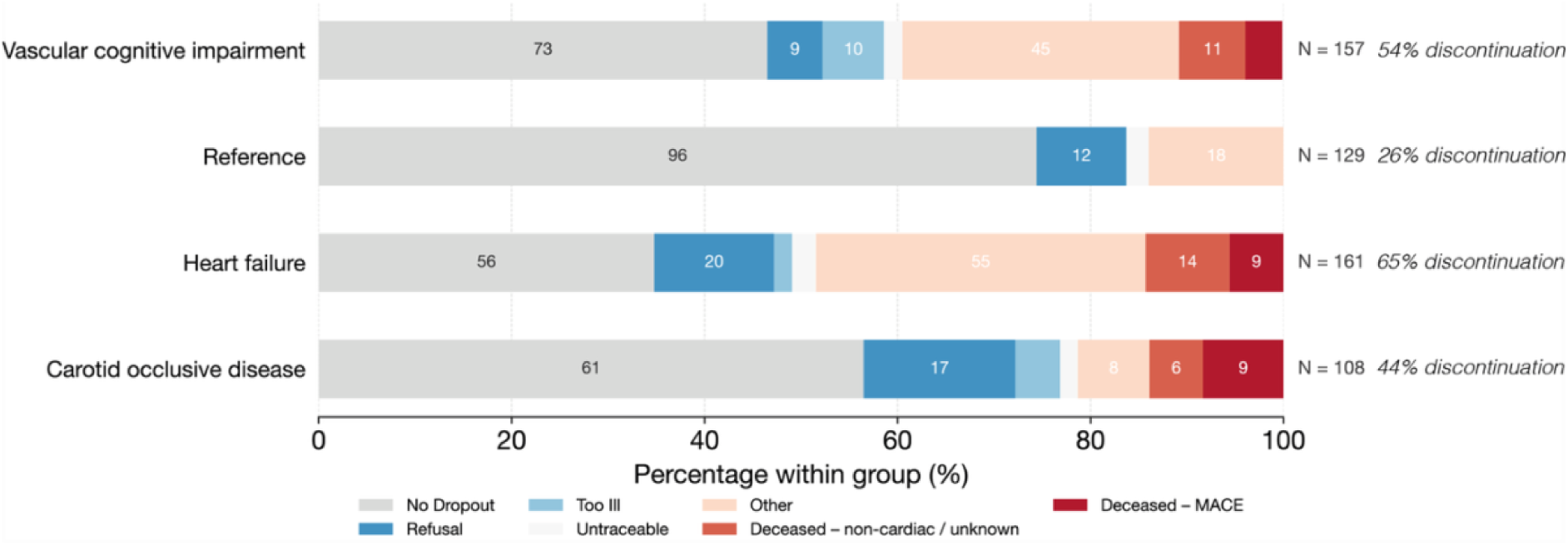
Reasons for study discontinuation per patient group.

**Table 1.** Baseline table.

| **n** | **566** |
| --- | --- |
| Age (median [IQR]) | 68.05 [61.58, 73.63] |
| Male sex (%) | 364 (64.3) |
| Diabetes (%) |  |
| Yes, with lifestyle advice | 16 (2.8) |
| Yes, with oral antidiabetics | 42 (7.5) |
| Yes, with insulin | 25 (4.4) |
| No | 480 (85.3) |
| Smoking (%) |  |
| Yes | 93 (16.5) |
| No | 150 (26.6) |
| Not anymore | 321 (56.9) |
| Systolic blood pressure (median [IQR]) | 140.50 [128.50, 154.50] |
| Low-density lipoprotein cholesterol (median [IQR]) | 2.60 [1.90, 3.30] |
| Systematic COronary Risk Evaluation 2 score (median [IQR]) | 8.30 [5.50, 12.97] |
| Blood pressure medication = No (%) | 201 (37.0) |
| Transient Ischemic Attack = No (%) | 423 (75.3) |
| Cerebrovascular Accident (%) |  |
| Yes, hemorrhage | 7 (1.2) |
| Yes, stroke | 122 (21.6) |
| Yes, type unknown | 3 (0.5) |
| No | 431 (76.1) |
| Patient group (%) |  |
| Carotid occlusive disease | 109 (19.3) |
| Heart failure | 162 (28.6) |
| Vascular cognitive impairment | 166 (29.3) |
| Reference | 129 (22.8) |

### Network structure

The constrained BN (Figure 2) shows a structure in which age and sex act as root nodes. Age shows high-frequency arcs to hippocampal/intracranial volume (% frequency over 200 bootstraps (f)= 99%) and the vascular risk score (f = 100%), and hippocampal/intracranial volume connects to brain/intracranial volume (f = 30%). The vascular risk score feeds into the SVD score (f = 78%), and the SVD score to patient group (f = 98%). Sex has arcs to stroke history (f = 10%) and atherosclerotic cardiovascular disease history (f = 83%), which in turn connect to patient group (f = 48% and f = 61%). Patient group strongly predicts baseline CDR (f = 100%), which has arcs to MMSE (f = 98%) and Starkstein score (f = 79%). The outcome MACE receives arcs from the MMSE score (f = 47%) and patient group (f = 89%), whereas CDR increase receives arcs from baseline CDR (f = 68%), and patient group (f = 100%).

**Figure 2.**
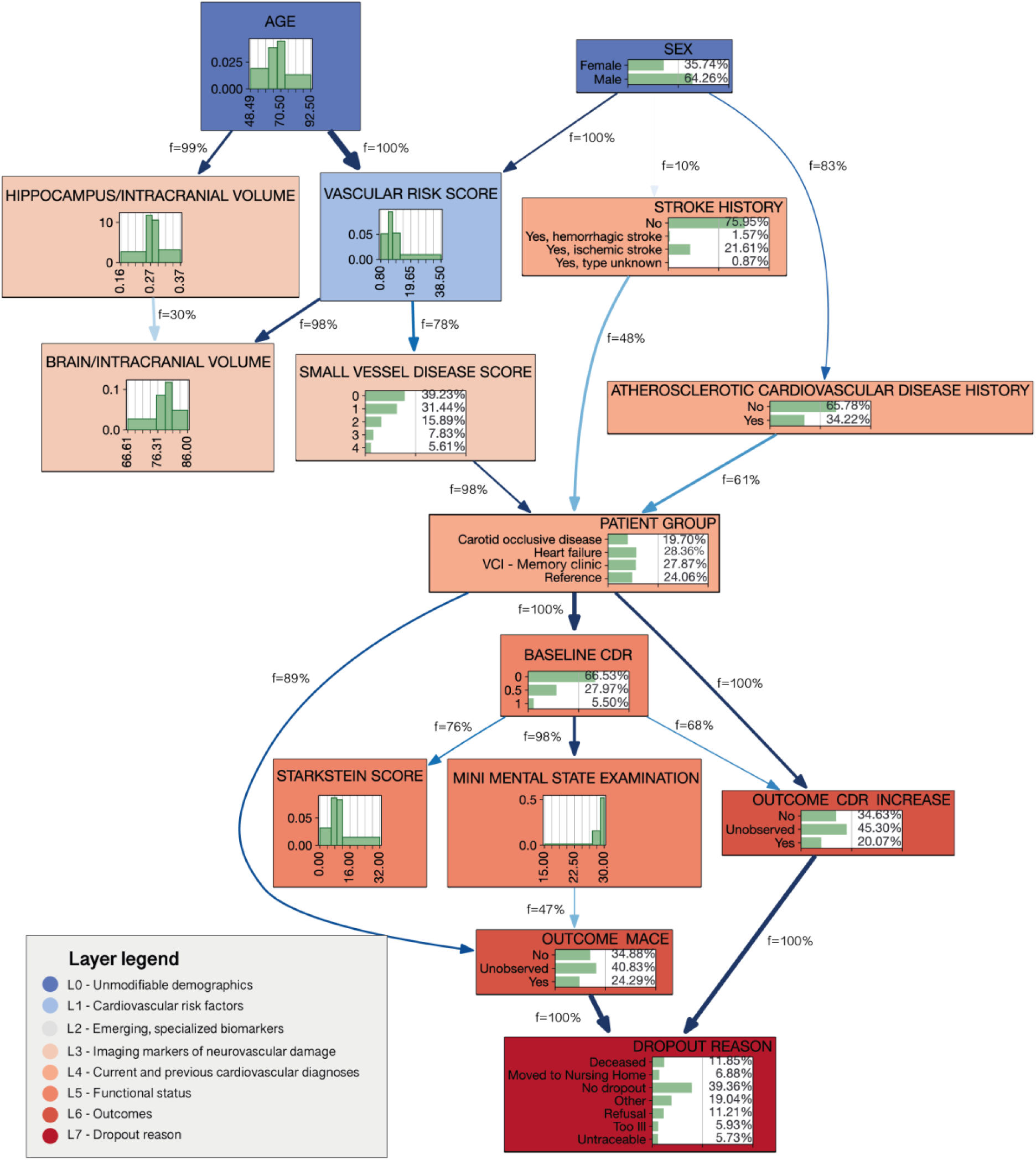
Clinically informed BN structure. *This figure shows the learned structure of the BN after applying a hierarchical, clinically informed variable order. Each box represents a variable, grouped by layer (L0–L7; shown at the bottom left). The arrows represent directed dependencies between variables within and between layers. Each arrow is accompanied by the bootstrap frequency (f = …%), which indicates the percentage of the 200 re-samples in which this relationship recurred; higher values indicate greater structural stability. Each box contains the empirical distribution of the variable (categorical or discretized). The outcome nodes (OUTCOME CDR INCREASE and OUTCOME MACE) and DROPOUT REASON are at the bottom and receive arrows from their direct predictors*.

For comparison, an unconstrained network was learned, in which no hierarchical layers were applied (Supplemental Figure S1). In this network, several arcs conflicted with known, plausible directions. For example, the vascular risk score appeared as a parent of age and sex. This implies that with the current sample size and number of factors included in the model, some degree of supervision of the model is required

### Jointly modelling cognitive decline, cardiovascular events, and dropout

The network jointly modelled interdependencies in factors related to the two clinical outcomes together with study dropout, and two patterns stood out from the conditional probability tables of the outcome nodes (Figure 2, Table 2). First, both observed cognitive decline and MACE are strongly related to baseline clinical status and diagnosis group (Figure 2). The main corresponding CPT, therefore, quantifies P(CDR outcome | baseline CDR, patient group) (Table 2). For participants with baseline CDR = 0, outcome probabilities differed markedly between patient groups. Reference participants had the highest probability of remaining cognitively stable, with P(CDR increase = No | Baseline CDR = 0, Reference) = 0.66, and a low probability of cognitive decline, P(CDR increase = Yes | Baseline CDR = 0, Reference) = 0.06. In contrast, memory-clinic patients with vascular brain lesions showed a substantially elevated probability of cognitive decline, with P(CDR increase = Yes | Baseline CDR = 0, Memory clinic–VCI) = 0.55. Patients with COD and HF occupied an intermediate position, with, at baseline, high probability for future dropout.

**Table 2.**
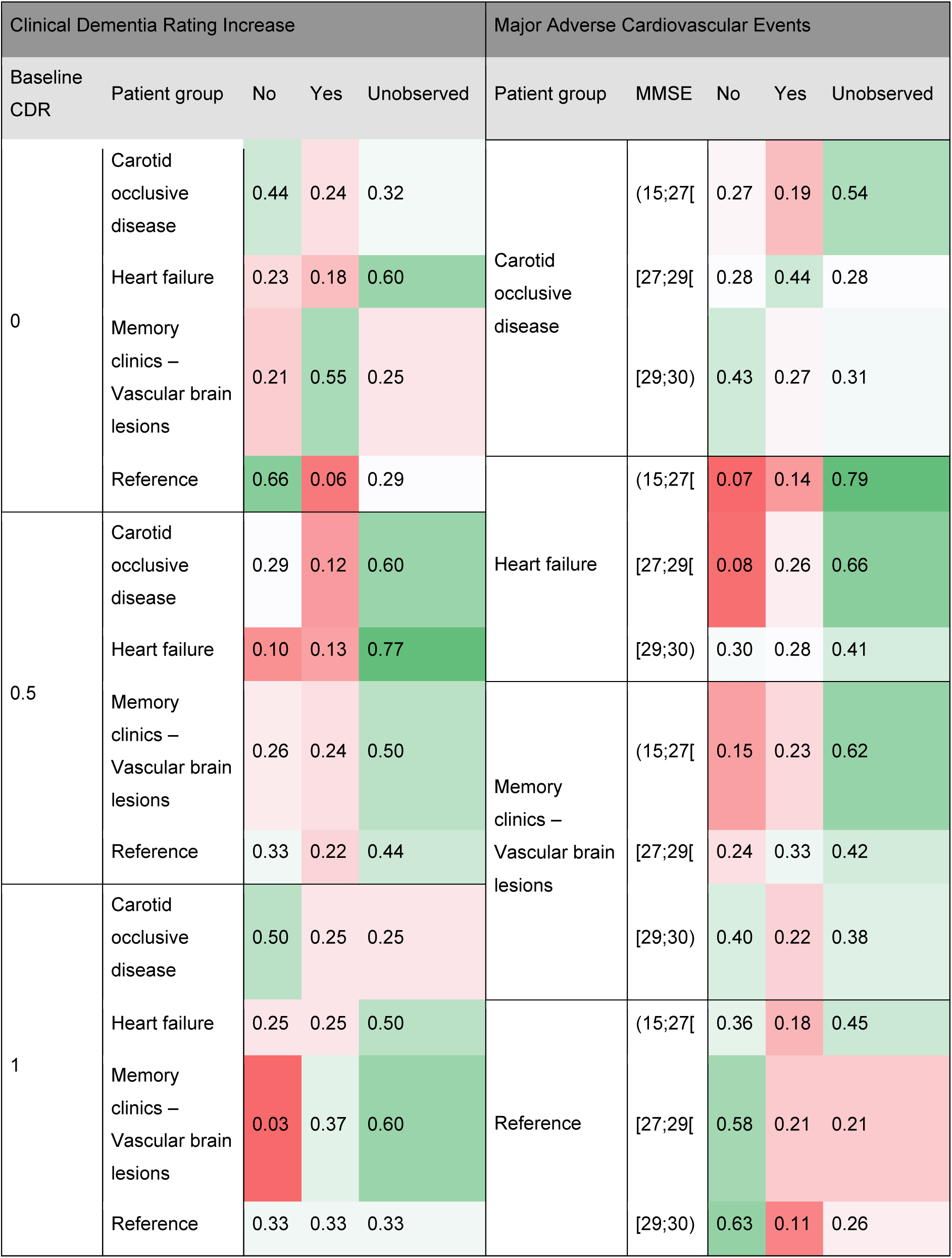
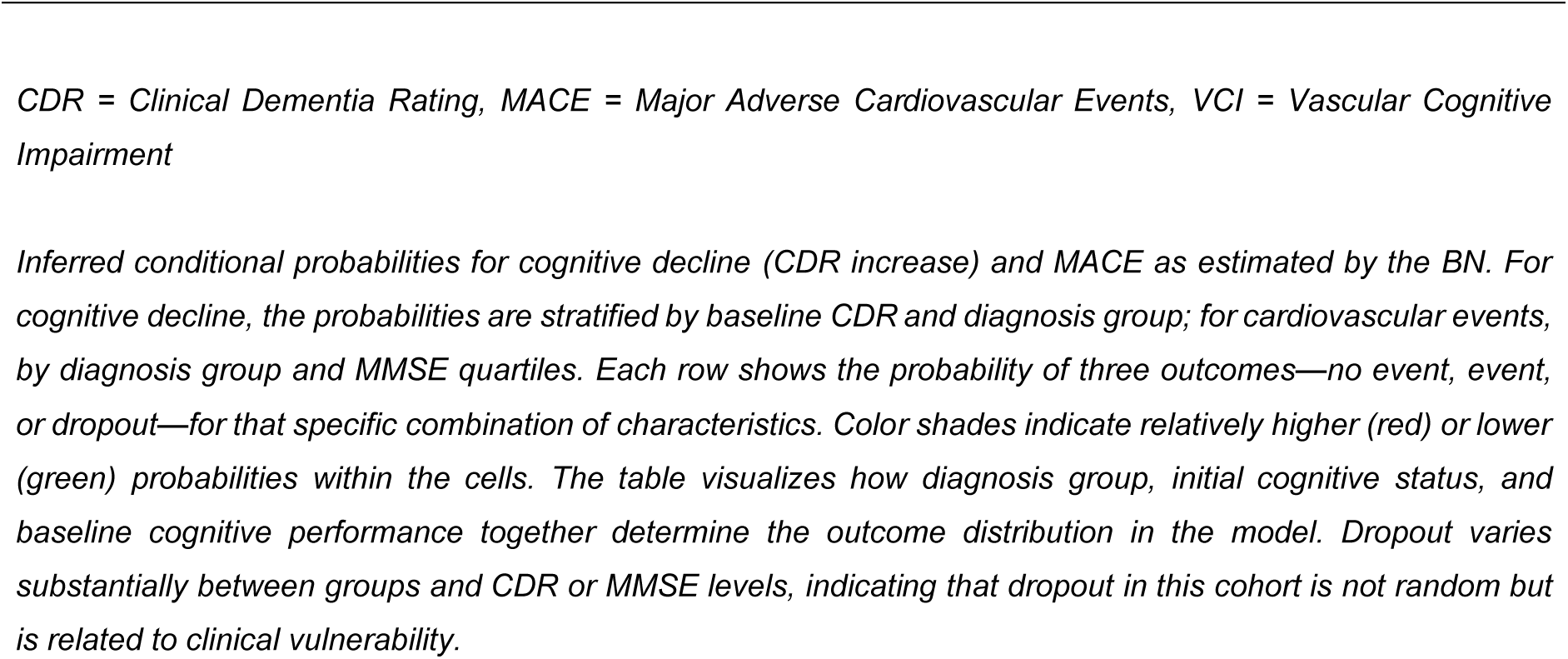
Conditional probability tables.

Second, worse cognitive functioning was related to more subsequent dropout, indicating that attrition in the cohort is related to clinical severity rather than random loss to follow-up (Table 2). At baseline CDR = 0.5, patients with HF showed the highest dropout probability, P(Unobserved | Baseline CDR = 0.5, HF) = 0.77, combined with low probabilities of no decline (P = 0.10) and CDR increase (P = 0.13) (Table 2). At this baseline level, probabilities for memory-clinic patients with vascular brain lesions were more evenly distributed across outcomes, whereas reference participants retained a comparatively lower dropout probability. At the highest baseline level (CDR = 1), outcome distributions diverged further by patient group. For reference participants, probabilities were approximately uniform across the three outcomes, with P(CDR increase = No | Baseline CDR = 1, Reference) ≈ P(CDR increase = Yes | Baseline CDR = 1, Reference) ≈ P(Unobserved | Baseline CDR = 1, Reference) ≈ 0.33, reflecting increased uncertainty in outcome prediction at advanced impairment. In contrast, memory-clinic patients with vascular brain lesions retained an elevated probability of cognitive decline, with P(CDR increase = Yes | Baseline CDR = 1, Memory clinic–VCI) = 0.37.

Similar patterns were observed for MACE, which was likewise modelled as a multinomial outcome (no observed event, observed event, unobserved), conditional on patient group and baseline MMSE category (Table 2). Among participants with COD and lower baseline cognitive performance (MMSE 15–27), the probability of an observed cardiovascular event was modest, P(MACE = Yes | COD, MMSE 15–27) = 0.19, whereas dropout was the most likely outcome, with P(Unobserved | COD, MMSE 15–27) = 0.54. In higher MMSE strata within this group, the probability of MACE increased, while dropout probabilities decreased accordingly. Across MMSE scores, patients with HF consistently showed high dropout probabilities, ranging from P(Unobserved | Heart failure, MMSE 15–27) = 0.79 to P(Unobserved | Heart failure, MMSE 29–30) = 0.41, with comparatively modest variation in the probability of observed cardiovascular events (P(MACE | Heart failure, MMSE 15–27) = 0.14 to P(MACE | Heart failure, MMSE 29–30) = 0.28).

In memory-clinic patients with vascular brain lesions, dropout probability declined with increasing baseline MMSE. Reference participants with high baseline cognitive performance (MMSE 29–30) showed the most favorable cardiovascular profile, with a low probability of MACE, P(MACE = Yes | Reference, MMSE 29–30) = 0.11, moderate dropout, P(Unobserved | Reference, MMSE 29–30) = 0.26, and the highest probability of remaining event-free, P(MACE = No | Reference, MMSE 29–30) = 0.63.

Taken together, dropout is a clinically informative outcome rather than random censoring, which has consequences for the interpretation of the other probabilities as well, meaning that the estimated probabilities for CDR increase and MACE are conditional on remaining under observation and may be biased if interpreted as unconditional (random-censoring) risks.

### Positioning of emerging imaging and plasma biomarkers within the network

When examining how emerging imaging and plasma biomarkers are positioned within the network and how they connect to established risk factors, imaging and outcomes, what stood out was that the biomarkers connected densely to each other but not to the outcome nodes (Figure 3). For example, NfL has outgoing arcs to GFAP (f = 50%), pTau181 (f = 100%), BCS-2 (f = 90%), and Aβ42 (f = 80%). However, the only outgoing arc from the biomarkers led to a history of atherosclerotic cardiovascular disease, with moderate bootstrap support (f = 20%). No other conditional dependencies were identified between the biomarkers and the downstream layers.

**Figure 3.**
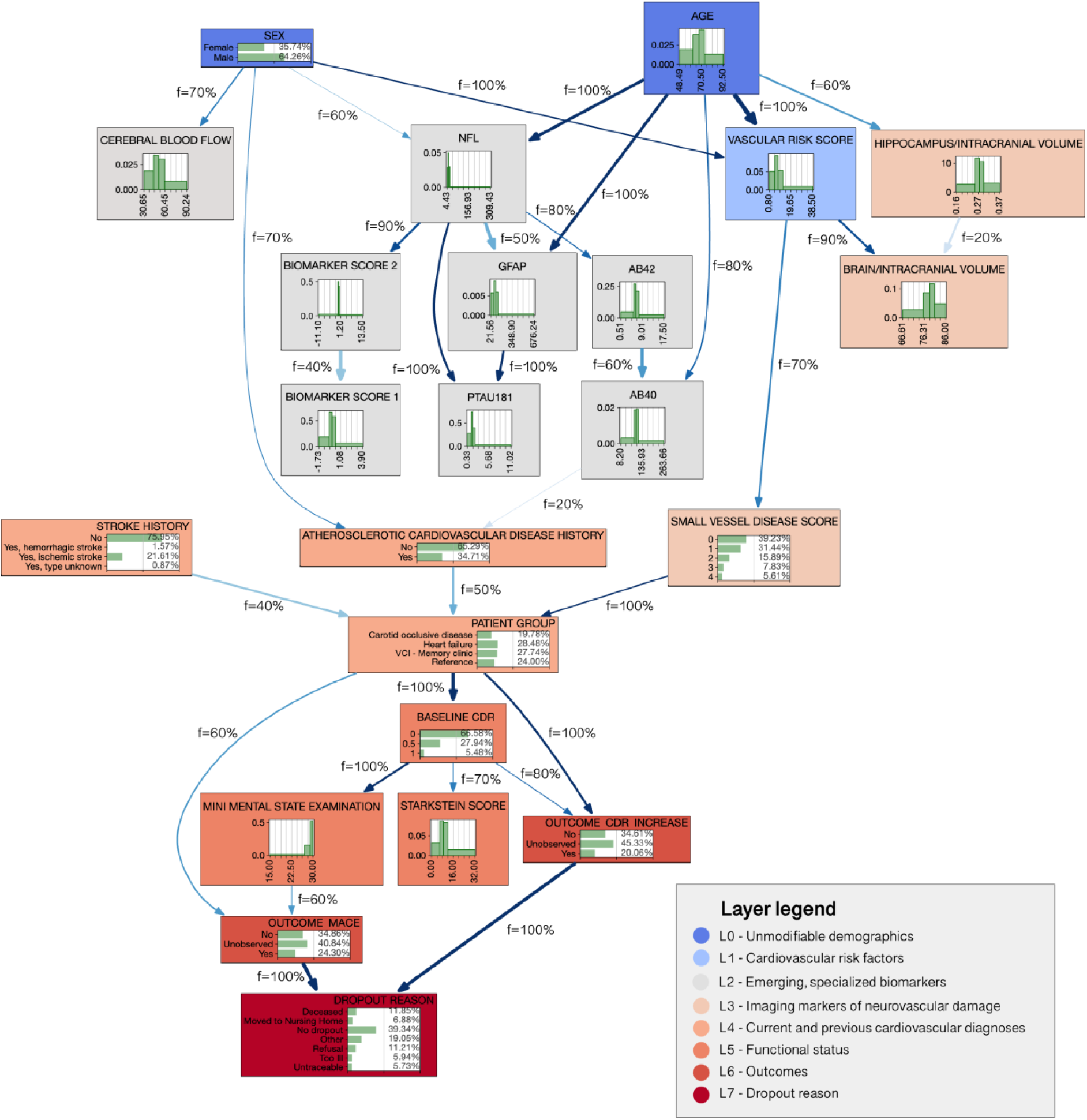
Extended BN structure including emerging biomarkers. *This figure shows the learned structure of the extended BN in which emerging blood- and CSF-based biomarkers were added as an additional layer. Variables are grouped by their predefined clinical layer (L0–L7; legend bottom right). Arrows represent directed dependencies learned from the data under the hierarchical constraints, and each arrow is annotated with its bootstrap frequency (f = …%), indicating how often the dependency recurred across 200 resamples. Higher frequencies denote greater structural stability. Each node displays the empirical distribution of that variable. Compared with the clinically constrained network without biomarkers, this extended structure shows how biomarkers connect upstream to demographic and vascular factors and to each other, but do not form stable direct arcs to the outcome nodes (OUTCOME CDR INCREASE and OUTCOME MACE)*.

For benchmarking, logistic regression analyses were performed for each biomarker in relation to both outcomes, unadjusted and adjusted for age and sex (Supplemental Table S1). Several plasma biomarkers showed statistically significant associations with observed cognitive decline and/or cardiovascular events in these models, for example, BCS-1 (OR = 2.58, 95% CI 1.71–3.90, p < 0.01) and GFAP (OR = 1.01, 95% CI 1.01–1.02, p < 0.01) for cognitive decline, and BCS-1 (OR = 1.68, 95% CI 1.20–2.35, p < 0.01) for MACE. After adjustment for age and sex, several associations persisted: BCS-1 (OR = 2.35, 95% CI 1.55–3.56, p < 0.01), BCS-2 (OR = 1.76, 95% CI 1.27–2.43, p < 0.01), GFAP (OR = 1.01, 95% CI 1.00–1.01, p < 0.01), and pTau181 (OR = 1.37, 95% CI 1.06–1.77, p = 0.02) for cognitive decline, and BCS-1 (OR = 1.48, 95% CI 1.03–2.11, p = 0.03) and BCS-2 (OR = 1.47, 95% CI 1.11–1.95, p < 0.01) for MACE. In the multivariate BN, however, no biomarker was connected to the outcome nodes by a direct arc. This pattern suggests that any biomarker–outcome association is weak and likely influenced by shared upstream factors such as vascular risk, SVD burden, and baseline cognitive status, rather than reflecting independent probabilistic pathways toward the outcomes.

### Probabilistic inference at the individual patient level

Lastly, we translated probabilistic inference to the individual patient level using a prototypical low-risk profile (58-year-old female, reference group, SVD score 0, baseline CDR = 0, MMSE = 29). The network assigned the highest probabilities to no CDR increase (mean 0.62, 95% CI 0.46–0.80) and no MACE (0.54, 0.40–0.67), with relatively low probabilities for observed CDR increase (0.10, 0.03–0.20) and observed cardiovascular events (0.18, 0.10–0.26) (Figure 4, Supplemental Table S2). To illustrate how the network updates probabilities as additional evidence is entered, we then fixed age (72 years) and sex (female) and varied the SVD score across its observed range (Figure 5). With increasing SVD score, the posterior probability of CDR increase rose modestly from 0.23 to 0.28, while the probability of MACE remained largely stable, illustrating that the responsiveness of the posterior depends on which factors are connected to the outcome in the learned structure.

**Figure 4.**
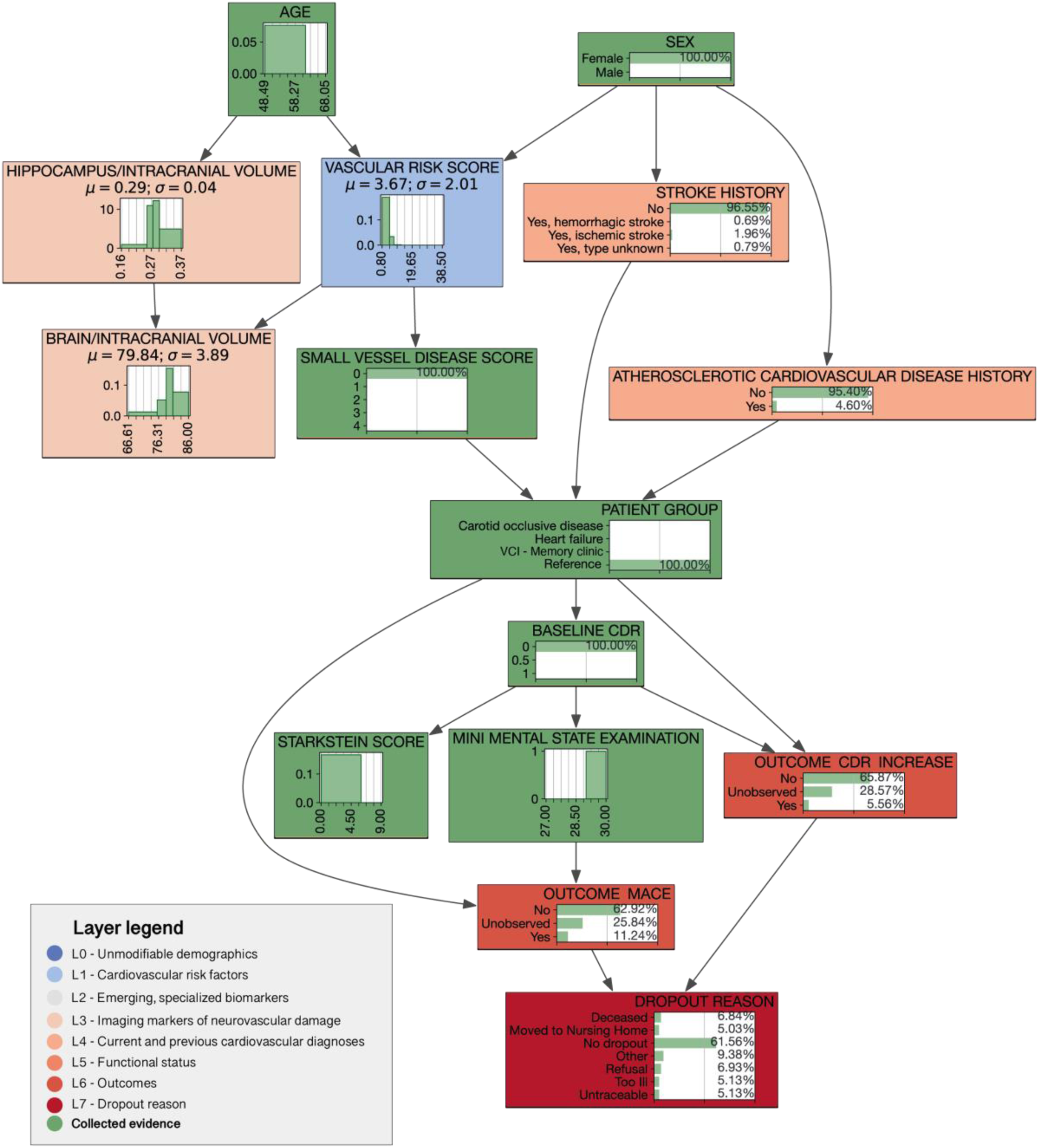
BN inference for an individual patient profile with collected evidence. *This figure illustrates how the BN updates posterior outcome probabilities when partial evidence is entered for a specific patient profile. Nodes shaded in green represent the variables for which evidence was provided (a 58-year-old female from the reference group, with an SVD score of 0, MMSE score of 29, and baseline CDR of 0). The grey arrows show the learned dependency structure, and unshaded nodes represent variables inferred rather than observed. Each node displays the posterior distribution given the entered evidence, allowing visualization of how information propagates through the network. The outcome nodes (OUTCOME CDR INCREASE, OUTCOME MACE, and DROPOUT REASON) reflect the individualized posterior probabilities resulting from this evidence propagation. This figure therefore shows how the joint structure of the model translates a limited set of patient characteristics into probabilistic predictions for multiple outcomes*.

**Figure 5.**
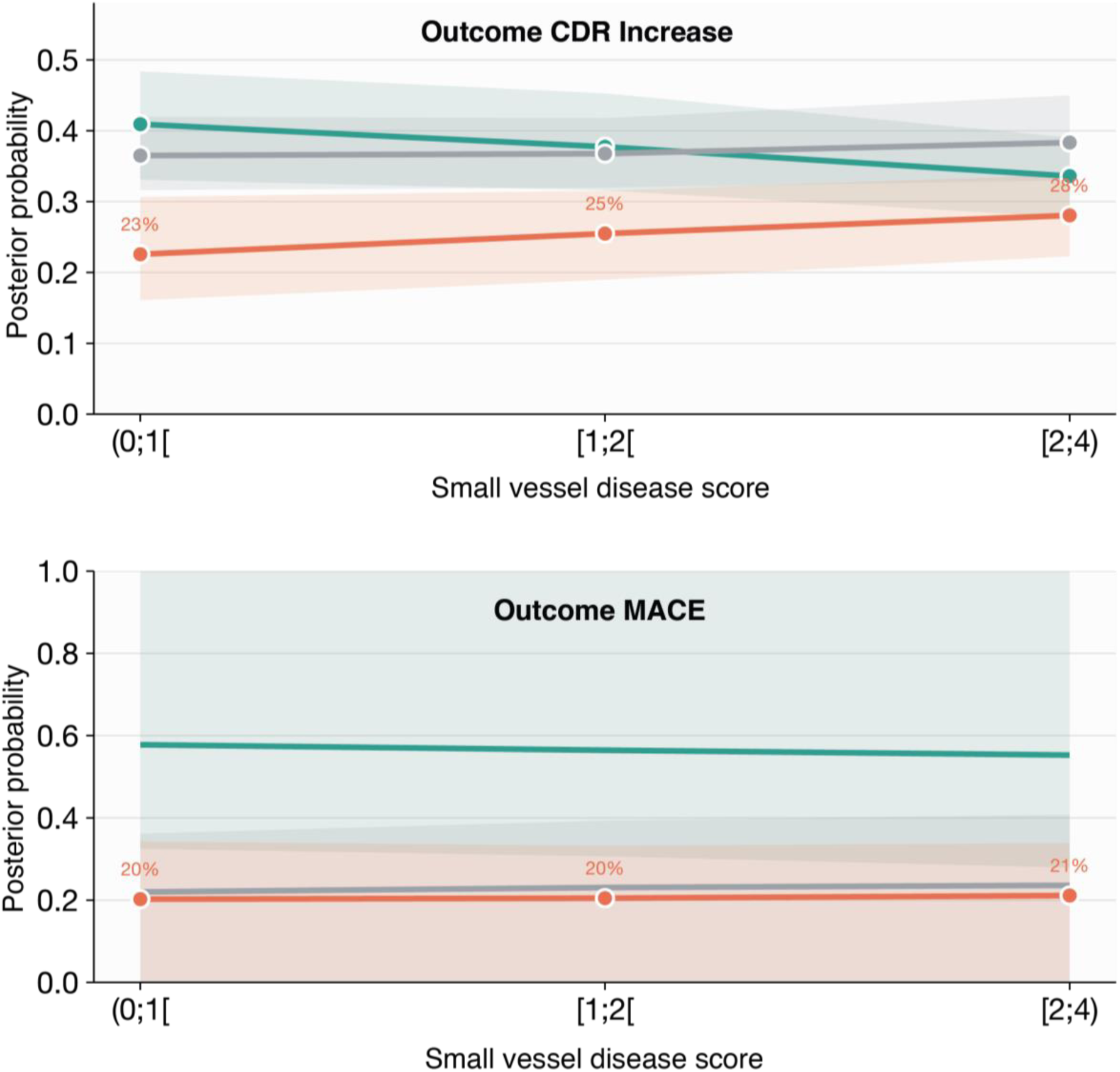
Updated outcome probabilities across small vessel disease scores for a 72-year old women. *This figure shows how the BN updates the posterior probabilities of both outcomes as the small vessel disease score increases and evidence (age 72 and female) is fixed. This illustrates how one or more of the factors in the network can be ‘tweaked’ and how the posterior probabilities are updated accordingly*.

## Discussion

In this study, we provide proof of concept of a BN framework to explore how vascular risk factors, neuroimaging markers, clinical characteristics and emerging plasma biomarkers jointly relate to cognitive decline, cardiovascular events, and dropout in a heterogeneous cohort of individuals with cardiovascular disease, a high burden of SVD, and at risk of VCI. The three complementary analyses illustrate that the method can provide insight in how: (i) combinations of baseline cognitive status and clinical condition strongly shaped the probabilities of both adverse outcomes and of selective attrition; (ii) emerging plasma biomarkers of neurodegenerative disease were not directly linked to outcomes in the network once vascular, imaging, and functional factors were included; and (iii) individualized posterior probabilities differed between prototypical low- and high-risk profiles, although bootstrap-based confidence intervals were wide relative to the differences between profiles. These findings thus show how probabilistic graph modelling can make the dependency structure underlying prognostic heterogeneity in this population explicit.

The application of BNs, in the context of VCI and SVD but also in other contexts, requires methodological decisions that shape the model structure and validity. Several key considerations warrant emphasis for researchers seeking to adopt this approach. First, the construction of a sparse, prior-knowledge based, layered network structure is nearly essential (26–28). In a BN, each additional variable and each extra category of that variable increases the number of probability values that must be estimated. When many variables with several categories are included, this quickly creates a very large table of possible combinations, most of which are almost never observed in the data. For example, if we model five variables with four categories each, there are already 4^5^ = 1,024 possible combinations, so in a dataset with 566 patients most combinations would be represented by only a number of patients. In such sparse tables, probability estimates become very unstable, and automated structure learning algorithms tend to pick up random patterns as if they were real, resulting in spurious connections and poor reproducibility across bootstrap samples 6/3/2026 11:20:00 AM. Our layered approach, sorting variables from demographics through vascular risk factors, imaging markers, and functional status to outcomes, serves multiple purposes: it reduces the computational complexity of the search space, prevents structurally implausible causal directions (such as outcomes influencing baseline characteristics, as shown in our results), and enhances clinical interpretability. While large-scale datasets with thousands of observations may accommodate less constrained structure learning, moderate sample sizes necessitate this sorting to avoid sparse CPT and unstable parameter estimates (29,30).

Second, discretization of continuous variables – as required by the method - has inherent trade-offs (31–33). Although discretization entails information loss, it enables the application of multinomial probability distributions, improves computational efficiency, and, when informed by clinical cut-points, enhances interpretability for decision-makers. In the standard BN framework employed in this study, we included no time component, as is the case with dynamic BNs. Unlike Cox proportional hazards or parametric survival models, static BNs do not directly accommodate censored time-to-event data or hazard ratios. While dynamic BN extensions exist for survival analysis, they introduce additional complexity and were not implemented here, but this could be an avenue for follow-up research (34,35). Beyond these methodological considerations, BNs offer several advantages for integrated prognostic modelling in heterogeneous cohorts. Unlike performing separate logistic regression analyses for each outcome, the BN simultaneously models cognitive decline, MACE, and attrition within a unified probabilistic framework. This integration provides three key benefits. First, it explicitly represents dropout as an informative outcome rather than treating it as random censoring or a nuisance variable, revealing that attrition in this cohort was strongly informed by baseline cognitive impairment and diagnostic group rather than occurring at random. Second, the CPT derived from the network offer a more dynamic and granular view of outcome probabilities than just baseline comparisons. For instance, the joint probability of cognitive decline for a memory clinic patient with baseline CDR = 0 differs markedly from that of a reference participant with identical cognitive status, information that would require multiple interaction terms in regression models. Third, when evaluating emerging plasma biomarkers, the BN approach facilitates assessment of added value by identifying whether plasma biomarkers exhibit direct probabilistic pathways to outcomes or whether their associations are influenced by established clinical, imaging, and functional variables.

In this cohort, logistic regression revealed statistically significant associations between several plasma biomarkers (BCS-1, BCS-2, GFAP, pTau181) and outcomes when adjusted only for age and sex (36). However, when embedded within the full BN, no biomarker formed stable direct arcs to outcome nodes. While BNs do not per se outperform logistic regression in predictive discrimination, they provide complementary mechanistic insights into the conditional dependency structure underlying prognostic heterogeneity, rendering explicit which combinations of patient characteristics jointly determine risk and which factors contribute unique information beyond established predictors. Overall, the modeling decisions, hierarchical layering, discretization strategies, and integrated multi-outcome representation, are methodologically more consequential than any single substantive finding, as they determine what questions the model can and cannot address and how reliably it can support clinical inference in populations at risk of VCI and SVD.

In relation to prior work, earlier applications of BNs in dementia have primarily addressed diagnostic differentiation using cognitive, clinical and structural imaging features rather than long-term prognosis, and mostly in the context of Alzheimer’s Disease (37–40). These studies showed that a BN can capture dependencies among cognitive and biological measures, but they were generally cross-sectional and did not incorporate vascular pathology, dropout mechanisms or multiple clinical outcomes. The present analysis extends this literature by applying a BN framework to a longitudinal, vascular-heterogeneous cohort and by jointly modelling cognitive decline, cardiovascular events and attrition.

Regarding validity and generalizability, a critical distinction must be made between internal structural robustness and external validation. By performing 200 bootstrap resamples, we ensured that the identified network structure and several dependencies are stable features of this dataset rather than artifacts of sampling noise. However, the specific conditional probabilities reported here are strictly cohort-specific, reflecting the recruitment settings and case-mix of the Heart-Brain Connection study, which applies to most studies. While we considered this cohort to be well suited to demonstrate the potential and pitfalls of BNs in VCI and SVD, the case mix clearly has limited external generalizability. The reader should therefore primarily regard the probability networks that were generated as an illustration of the methodological framework rather than precise risk estimates. Nonetheless, the hierarchical structure learning pipeline is transferable and can be applied to independent cohorts to derive site-specific dependency maps, prioritizing methodological reproducibility over result reproducibility. Future work incorporating dynamic BNs to evolve over time, continuous-variable methods, and external replication is warranted.

In summary, this study provides a methodological proof-of-concept for using BNs to organize multimodal data in an heterogeneous VCI and SVD cohort. While the specific risk probabilities presented reflect the unique characteristics of our population, the approach successfully showcases how vascular, imaging, and functional factors jointly organize to influence outcomes and selective attrition. By embedding emerging imaging and plasma biomarkers within this broader dependency structure, we demonstrated a transparent method for evaluating their incremental value beyond established clinical predictors. We advocate the use of frameworks like this to offer researchers an analytical approach to visualize complex, conditional dependency structures underlying prognosis in multi-morbid patient populations.

## Acknowledgements

We gratefully acknowledge the contribution of researchers and participants of the HBCx consortium.

## Author Contributions

**Malin Overmars:** Conceptualization, Formal analysis, Methodology, Software, Validation, Visualization, Writing – Original Draft, Writing – Review & Editing. **Cor Allaart**: Writing – Review & Editing, Investigation, Funding acquisition. **Esther Bron**: Writing – Review & Editing, Data Curation, Formal Analysis, Funding acquisition, Resources, Investigation. **Hans-Peter Brunner La Rocca**: Writing – Review & Editing, Investigation, Funding acquisition. **Jeroen de Bresser:** Writing – Review & Editing, Resources, Investigation, Data Curation, Funding acquisition. **Majon Muller:** Writing – Review & Editing, Investigation, Funding acquisition. **Matthias van Osch**: Writing – Review & Editing, Resources, Investigation, Data Curation, Funding acquisition. **Charlotte Teunissen**: Writing – Review & Editing, Resources, Investigation, Data Curation, Funding acquisition. **Betty Tijms:** Writing – Review & Editing. **Frank Wolters** Writing – Review & Editing. **Geert Jan Biessels**: Conceptualization, Funding acquisition, Investigation, Supervision, Writing – Original Draft, Writing – Review & Editing

## Statements and Declarations

### Ethical considerations

The protocol received central approval from the Medical Ethics Review Committee of Leiden University Medical Center, and local approval was obtained at each participating site. The Heart-Brain Study was conducted in accordance with the Declaration of Helsinki (2013 version) and the Dutch Medical Research Involving Human Subjects Act (WMO).

### Consent to participate

All participants provided written informed consent prior to any study-related procedures.

### Data Availability Statement

All code used for data processing, BN modelling and analysis is publicly available at: https://github.com/umcu/vci-bayes. The Heart–Brain Connection data are not publicly available due to privacy regulations but can be requested from the corresponding author upon reasonable request and with appropriate approvals.

