## Supplemental Methods for "Bayesian networks to estimate prognosis in vascular cognitive impairment and small vessel disease: integrated analyses of interdependent contributors to multiple outcomes"

**Supplemental Material**

**Supplemental Figures**

**Supplemental Figure S1.** Unconstrained BN structure learned without clinically informed hierarchical ordering

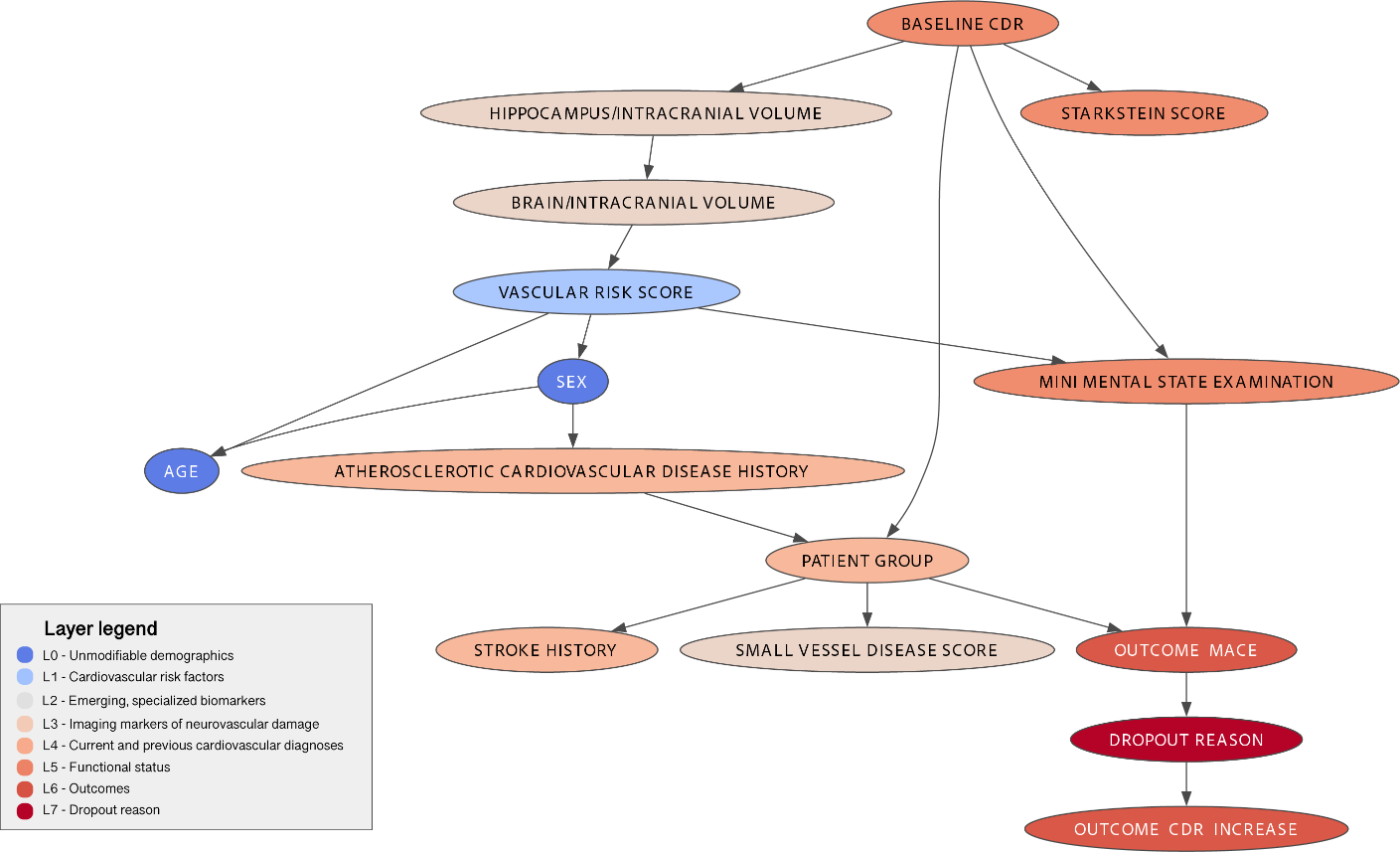

*This visualization shows the structure of the unrestricted BN, in which no clinically informed hierarchical ordering has been set. The arrows represent directed dependencies learned by the algorithm without restrictions on possible directions. Variables are grouped according to the same color layers (L0–L7) as in the constrained models, but the relationships shown are derived solely from the data. In this network, various connections arise that do not correspond to known or plausible clinical directions. For example, the vascular risk score appears as a parent variable of age and sex, even though this score is partly determined by age and sex, and there is an arrow from the baseline CDR to structural brain volumes, indicating reverse causality. The figure thus shows how, in the absence of clinical structural constraints, the model learns dependencies that are statistically possible but not interpretable in terms of content or causally implausible. This network therefore serves as a reference point.*

**Supplemental Tables**

**Supplemental Table S1**. Unadjusted and age- and sex-adjusted logistic regression associations between emerging plasma biomarkers and five-year cognitive decline (CDR increase) and major adverse cardiovascular events

| **Outcome** | **Biomarker** | **N** | **Odds Ratio (95% CI) (Unadjusted)** | **Odds Ratio (95% CI) (Adjusted age + sex)** | **P-value (Unadjusted)** | **P-value (Adjusted age + sex)** |
| --- | --- | --- | --- | --- | --- | --- |
| CDR increase | Aβ40 | 304 | 1.01 (1.00, 1.03) | 1.01 (1.00, 1.02) | 0.02 | 0.14 |
| CDR increase | Aβ42 | 304 | 1.09 (0.96, 1.24) | 1.07 (0.94, 1.22) | 0.18 | 0.28 |
| CDR increase | BCS-1 | 304 | 2.58 (1.71, 3.90) | 2.35 (1.55, 3.56) | < 0.01 | < 0.01 |
| CDR increase | BCS-2 | 304 | 1.84 (1.36, 2.51) | 1.76 (1.27, 2.43) | < 0.01 | < 0.01 |
| CDR increase | CBF | 304 | 0.98 (0.96, 1.00) | 0.99 (0.97, 1.01) | 0.10 | 0.33 |
| CDR increase | GFAP | 304 | 1.01 (1.01, 1.02) | 1.01 (1.00, 1.01) | < 0.01 | < 0.01 |
| CDR increase | NfL | 304 | 1.01 (1.00, 1.03) | 1.01 (1.00, 1.02) | 0.07 | 0.24 |
| CDR increase | pTau181 | 304 | 1.53 (1.19, 1.98) | 1.37 (1.06, 1.77) | < 0.01 | 0.02 |
| MACE | Aβ40 | 328 | 1.00 (0.99, 1.01) | 0.99 (0.98, 1.00) | 0.99 | 0.17 |
| MACE | Aβ42 | 328 | 0.92 (0.82, 1.04) | 0.90 (0.80, 1.02) | 0.17 | 0.10 |
| MACE | BCS-1 | 328 | 1.68 (1.20, 2.35) | 1.48 (1.03, 2.11) | < 0.01 | 0.03 |
| MACE | BCS-2 | 328 | 1.68 (1.29, 2.21) | 1.47 (1.11, 1.95) | < 0.01 | < 0.01 |
| MACE | CBF | 328 | 0.98 (0.96, 1.00) | 0.99 (0.97, 1.01) | 0.02 | 0.34 |
| MACE | GFAP | 328 | 1.00 (1.00, 1.01) | 1.00 (1.00, 1.01) | 0.03 | 0.75 |
| MACE | NfL | 328 | 1.02 (1.00, 1.04) | 1.01 (0.99, 1.02) | 0.01 | 0.26 |
| MACE | pTau181 | 328 | 1.02 (0.83, 1.27) | 0.83 (0.65, 1.06) | 0.84 | 0.14 |

**Supplemental Table S2**. Bootstrap-derived posterior probability distributions for cognitive decline and major adverse cardiovascular events for a prototypical low-risk patient profile inferred from 200 resampled Bayesian networks

| **Scenario** | **Outcome** | **Category** | **Mean probability** | **Standard deviation** | **CI 2.5%** | **CI 97.5%** |
| --- | --- | --- | --- | --- | --- | --- |
| Age 58, Female sex, Reference, SVD-score = 0, MMSE = 29, Baseline CDR = 0 | CDR increase | No | 0.62 | 0.08 | 0.46 | 0.80 |
| Age 58, Female sex, Reference, SVD-score = 0, MMSE = 29, Baseline CDR = 0 | CDR increase | Unobserved | 0.28 | 0.07 | 0.14 | 0.38 |
| Age 58, Female sex, Reference, SVD-score = 0, MMSE = 29, Baseline CDR = 0 | CDR increase | Yes | 0.10 | 0.05 | 0.03 | 0.20 |
| Age 58, Female sex, Reference, SVD-score = 0, MMSE = 29, Baseline CDR = 0 | MACE | No | 0.54 | 0.08 | 0.40 | 0.67 |
| Age 58, Female sex, Reference, SVD-score = 0, MMSE = 29, Baseline CDR = 0 | MACE | Unobserved | 0.28 | 0.05 | 0.20 | 0.38 |
| Age 58, Female sex, Reference, SVD-score = 0, MMSE = 29, Baseline CDR = 0 | MACE | Yes | 0.18 | 0.05 | 0.10 | 0.26 |

**Supplementary Methods**

**MRI processing**

Processing of the brain MRI was performed using two automated pipelines. For each patient, manual segmentation of infarcts and other pathologies that potentially affect automatic tissue segmentation was performed by a neuroradiologist. Subsequently, the annotated infarcts and pathologies were manually segmented by trained students. In addition, an automated pipeline (Quantib Brain, Rotterdam, the Netherlands) was used to segment WMH based on FLAIR scans. A brain tissue segmentation method was applied to the 3D T1-weighted images. From these segmentations, volumes in milliliters (mL) of total brain gray matter (GM), white matter, cerebrospinal fluid, and WMH were computed.

**Plasma biomarkers**

Ethylenediaminetetraacetic acid (EDTA) plasma was obtained by venipuncture and, after centrifugation at 1,800×g for 10 minutes at room temperature, aliquoted (0.5 mL) into polypropylene tubes and stored at −80°C. Before analysis, samples were briefly thawed at room temperature and spun at 10,000×g for 10 minutes to remove residual debris that could interfere with the assays. Plasma Aβ42, Aβ40, GFAP, and NfL concentrations were quantified using the Simoa Neurology 4‑plex E kit (Quanterix, Billerica, MA, USA), and plasma pTau181 was measured with the Simoa pTau181 V2 kit (Quanterix) on the Simoa HD‑X platform. All measurements were performed in duplicate and followed the manufacturer’s protocol, including an automated 1:4 on‑board dilution step. The mean intra‑assay coefficients of variation (CVs) were 2.4% for Aβ40, 2.6% for Aβ42, 5.1% for GFAP, 4.3% for NfL, and 7.4% for pTau181, while inter‑assay CVs were 6.5% for Aβ40, 5.9% for Aβ42, 6.7% for GFAP, and 5.7% for NfL, based on three quality‑control samples across seven runs; for pTau181, the mean inter‑assay CV was 7.8%, calculated from two quality‑control samples measured in six runs. Two previously published Olink plasma biomarker compound scores (BCS), reflecting inflammation and coagulation processes, have also been added, which are described in more detail elsewhere (19,20).

**Cerebral blood flow**

Normal appearing gray matter CBF was measured with pCASL (multi-slice 2D echo-planar imaging [EPI] acquisition with background suppression; labeling duration = 1800 milliseconds; post-labeling delay = 1800 milliseconds; single-shot EPI readout; resolution = 3 × 3 ×7 mm3).2,12 pCASL data were processed using the automated Iris pipeline for CBF quantification . Quantification of pCASL data into CBF maps was based on a single- compartment model after the subtraction of labeled images from control images.12 To scale the signal intensities of the subtracted pCASL images to absolute CBF units, a separately acquired proton density weighted image was used. The quantification further included motion-correction of the raw pCASL data and partial volume correction (PVC). CBF was quantified in normal-appearing gray matter only. To obtain the normal-appearing gray matter mask for each participant, first a binary gray matter segmentation was obtained. Subsequently, PVC-uncorrected pCASL images of all participants were visually inspected. Images with suboptimal quality (i.e., motion artefacts, incomplete ASL-sequence, or labeling errors) and images with dominant vascular artefacts and little tissue perfusion signal were not used for further analyses.
